# Protocol: WorkoutCPP - a pilot series of N-of-1 trials evaluating RL-generated adaptive exercise recommendations for pelvic pain management

**DOI:** 10.64898/2026.09.25.26356699

**Authors:** Bryan T. Tricoche, Stefan Konigorski, Rhyann Clarke, Johannes E. Vedder, Matteo Danieletto, Kyle Landell, Jovita Rodrigues, Alexander Kim, Gerard Ona, Christine Olivo, Andrew Dorsey, Sharonne Holtzman, Susan Khalil, Ipek Ensari

**Author notes:** Corresponding author: Ipek Ensari, PhD, Windreich Department of Artificial Intelligence and Human Health, Icahn School of Medicine at Mount Sinai, 3 E 101^st^ Street, Room 1009, New York, NY, 10029. Trial Sponsor: Icahn School of Medicine at Mount Sinai, One Gustave L. Levy Place, New York, NY 10029. The sponsor had no role in study design, data collection, analysis, interpretation, or the decision to submit for publication. Funding was provided by the Hasso Plattner Institute Transatlantic Pilot Award (co-PIs: Ensari, Konigorski). Study oversight is provided by the PI (I.E.) in consultation with the co-investigator (S.K.); no independent steering committee has been established given the pilot scope and minimal risk profile of the study.

## Abstract

**Introduction:** Chronic pelvic pain disorders (CPPDs) affect up to one in four reproductive-age women and are characterized by fluctuating, heterogeneous symptoms that complicate engagement in regular physical exercise. Standard programs rarely adjust to daily symptom variability. *WorkoutCPP* is a pilot series of N-of-1 trials evaluating the feasibility of an adaptive mobile-health (mHealth) intervention using reinforcement learning (RL) to personalize exercise for individuals with CPPDs.

**Methods and Analysis:** This pilot study consists of a series of N-of-1 trials employing a randomized ABAB or BABA design, in which 45 participants (aged 18-55 years) with CPPDs each serve as their own control. Each participant completes one baseline week followed by four alternating 2-week intervention phases, totaling 9 weeks, comparing an active comparator arm (A = generic exercise guidance) and an experimental arm (B = adaptive RL-generated recommendations). The RL agent integrated into the ehive app suggests daily exercise recommendations based on participant-reported pain and prior exercise behavior. Primary outcomes are exercise recommendation adherence rate and participant retention rate; safety and additional feasibility indicators are also monitored. Secondary outcomes assess algorithm interpretability. Exploratory outcomes assess contextual sensitivity of the RL algorithm and changes in pain and behavioral engagement.

**Ethics and Dissemination:** Ethical approval was granted by the Icahn School of Medicine IRB (Protocol # STUDY-23-00721). Electronic informed consent will be obtained. Findings will be shared via peer-reviewed publications, conferences, and de-identified data summaries.

**Registration:** This study protocol was publicly registered on the Open Science Framework (OSF) Registries (URL: https://osf.io/d9f36) prior to participant enrollment, to support international open-science data transparency and reproducibility standards for mobile health adaptive intervention trials. The study was subsequently registered on ClinicalTrials.gov (NCT07810218), consistent with ICMJE recommendations for trials informing future confirmatory research. All documented study procedures match the baseline human subjects tracking mechanisms approved by the Icahn School of Medicine at Mount Sinai Institutional Review Board (Protocol # STUDY-23-00721).

**Article Summary:** *Strengths and Limitations:*

1. This study represents the first application of reinforcement-learning–based adaptive exercise recommendations for individuals with chronic pelvic pain disorders.
2. N-of-1 trial framework enables detailed within-person assessment of feasibility and individual responses.
3. Integration of wearable and app-based data allows real-time contextual adaptation.
4. While the ABAB/BABA design supports within-person comparisons, variability across participants (person-level differences) may limit the generalizability of findings.
5. As a pilot study, the sample is small, findings are exploratory and intended to inform the design of future confirmatory trials.
6. Although participants are blinded to their assigned arm at any given time, they may infer which study arm they are part of based on the nature of the recommendations.

## Introduction

Chronic pelvic pain disorders (CPPDs) comprise a spectrum of gynecologic, urologic, gastrointestinal, and musculoskeletal conditions that persist for at least three to six months and frequently overlap in presentation, as reflected in recent classification frameworks.(1-3) It is estimated that between 16– 25% of reproductive-age women are affected by CPPDs, such as endometriosis, adenomyosis, and fibroids.(2) They impose a public health burden through diminished quality of life (QoL), reduced productivity, sexual dysfunction, and high healthcare utilization.(4, 5) Despite their prevalence, CPPDs remain under-recognized and fragmented across medical disciplines, often leading to diagnostic delays and insufficient care.(5) CPPDs often involve chronic overlapping pain conditions with multiple contributing pain generators, including organ-based and non-organ-based mechanisms. Management typically requires multidisciplinary approaches, with non-surgical and non-pharmacological strategies serving as important adjuncts to improve quality of life.

CPPDs are multifaceted and heterogeneous in their causes, manifestations, and impact. Their etiology can involve hormonal, inflammatory, and neuromuscular mechanisms that interact in complex and individualized ways.(1) Symptom patterns vary not only between individuals but also within individuals over time, fluctuating with stress, hormonal cycles, activity levels, and sleep.(2, 6) This heterogeneity extends beyond pain intensity to include fatigue, mood disturbances, and pain self-efficacy, creating unique challenges for designing consistent, effective behavioral interventions.(6) As a result, therapeutic responses are highly variable, and standard exercise or rehabilitation programs often fail to meet patients’ day-to-day needs and fluctuations in symptoms.(6, 7)

Physical activity (PA), particularly structured and goal-oriented exercise, is emerging as a promising, low-risk approach for symptom management in chronic pelvic pain and related conditions.(8, 9) Preliminary studies in broader chronic pain populations suggest that gentle or graded exercise can improve pain interference, fatigue, and overall physical function.(6, 10) However, sustaining consistent participation represents a challenge for individuals with CPPDs, as symptoms often fluctuate from day to day.(6) Uncertainty about appropriate activity intensity or timing during pain flares may contribute to variable adherence and limited long-term engagement. Approaches that allow exercise intensity and type to adapt dynamically to symptom state could better support safety and participation over time.

Digital and mobile health (mHealth) platforms offer a scalable means to deliver adaptive and personalized guidance. Most commercially available applications, however, still rely on static recommendations or generic reminders that do not reflect the complexity or variability of CPPD symptoms.(6, 11, 12) Reinforcement learning (RL), a branch of machine learning, offers a dynamic framework for adaptive decision-making by continuously learning which actions maximize desired outcomes through iterative interaction and feedback.(13) Our team previously developed the *ehive* application (14), a centralized mHealth research infrastructure that enables multimodal data collection from user-tracking and wearables. It has been used for observational studies enrolling individuals with CPPDs (14) to collect participant-tracked and device-based data on health status and behaviors.

Leveraging this infrastructure, the *WorkoutCPP* study applies RL-based adaptive recommendation methods to personalize daily activity suggestions according to each participant’s evolving pain, fatigue, and behavioral context, with safety maintained through predefined constraints. Generic, evidence-based exercise recommendations were selected as the active comparator arm because they represent standard, non-personalized care currently available to individuals with CPPDs, providing a meaningful baseline against which to evaluate the added value of RL-based personalization.

Individuals with CPPDs experience highly variable symptom patterns and may have varied responses to PA interventions, making evaluation of treatment effects at the individual level critical for understanding which strategies are effective for whom.(4) N-of-1 trials provide a rigorous framework for evaluating individualized behavioral interventions by allowing each participant to serve as their own control across alternating intervention arms.(12, 15) This approach is particularly well suited to CPPDs, which are characterized by both substantial within-person symptom fluctuations and heterogeneity in responses between individuals, limiting the interpretability of group-level treatment effects.

A series of N-of-1 trials enables systematic within-person comparisons while still allowing aggregation of findings across participants to inform broader intervention development. Existing digital and mHealth tools for pain typically offer static or generalized guidance, and RL approaches have not been extensively evaluated using real-world data implemented for individuals living with CPPDs in diverse populations.(12, 14, 15) We previously demonstrated the feasibility of an online RL agent that generated adaptive exercise recommendations using a N-of-1 trial framework for endometriosis-related pain management (12), providing an initial proof-of-concept for integrating RL into personalized exercise research. Whereas that study primarily established algorithmic feasibility in a limited sample, WorkoutCPP extends this work by investigating real-world clinical implementation and evaluating feasibility, safety, and model interpretability in a prospective pilot trial.

Accordingly, we herein extend prior proof-of-concept findings to evaluate whether adaptive RL-driven exercise recommendations can be safely and feasibly implemented among individuals with CPPDs. Specifically, this pilot study will:

1. Evaluate participant engagement and safety during daily adaptive exercise recommendations delivered via the ehive mHealth platform.
2. Assess study design feasibility and data quality, including recruitment, retention, adherence, and completeness of repeated daily and weekly measures within the N-of-1 ABAB/BABA framework.
3. Validate and interpret the reinforcement-learning (RL) model, examining how contextual factors influence action probabilities and how safety-based fallback logic operates in practice.

The investigation of these pilot aims will inform the refinement of the RL algorithm and the design of a larger randomized controlled trial evaluating the efficacy and scalability of personalized, data-driven exercise interventions for CPPDs.

## Methods

### Study design and setting

This pilot N-of-1 intervention study aims to evaluate the feasibility, safety, and preliminary effects of adaptive exercise interventions for individuals with chronic pelvic pain disorders (CPPDs). Participant recruitment began on February 6, 2026, and is ongoing, with an anticipated study completion date of August 6, 2027. Each participant will serve as their own control, completing four alternating 2-week phases following a 1-week baseline period. Participants will be randomized to one of two sequences, ABAB or BABA. Participants are blinded to their assigned arm. The two arms are as follows:

- **Active Comparator arm (A):** generic evidence-based exercise recommendations, serving as the control condition for this study, and
- **Experimental arm (B):** adaptive exercise recommendations generated by a reinforcement-learning (RL) agent.

Both arms are delivered in the same format, a daily notification specifying an activity, duration, and intensity, helping preserve participant blinding to arm assignment.

A one-week baseline run-in period will precede randomization to allow participants to become familiar with the ehive app and to initialize algorithm parameters. Participants may extend their study period by up to two additional weeks (11 weeks total) to accommodate missing data or technical difficulties. The default total participation period is nine weeks (one week baseline plus eight weeks of alternating intervention phases) for which a timeline is shown in Figure 1.

**Figure 1.**
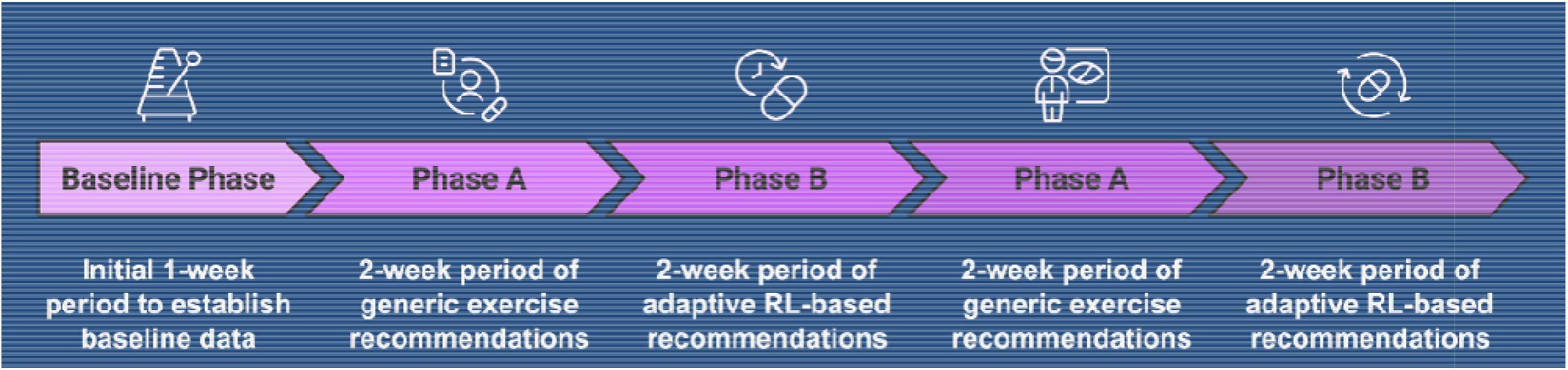
Schematic overview of the WorkoutCPP N-of-1 trial design. Each participant completes a 1-week baseline period followed by four alternating 2-week intervention phases. Participants randomized to the ABAB sequence receive the active comparator arm (generic exercise recommendations) followed by the experimental arm (adaptive RL-generated recommendations), repeated twice. Participants randomized to the BABA sequence complete the arms in reverse order. Participants may extend up to 11 weeks total under the conditions described in the main text, but this extension is not pictured in this figure.

Exercise recommendations will be delivered once daily through the ehive platform. The daily cadence is intended to maximize opportunities for engagement and adherence while accommodating fluctuations in pain, fatigue, or schedule. Participants are not expected to complete every recommendation; rather, the RL agent learns future recommendations based on completion patterns and self-reported responses. This higher-frequency, low-burden approach supports sustained engagement (16) and enables the collection of richer contextual data, which is essential for individualized adaptation in adaptive mHealth interventions (11). A schematic overview of the system architecture and application interface are provided in Figures 2 and 3, respectively.

**Figure 2.**
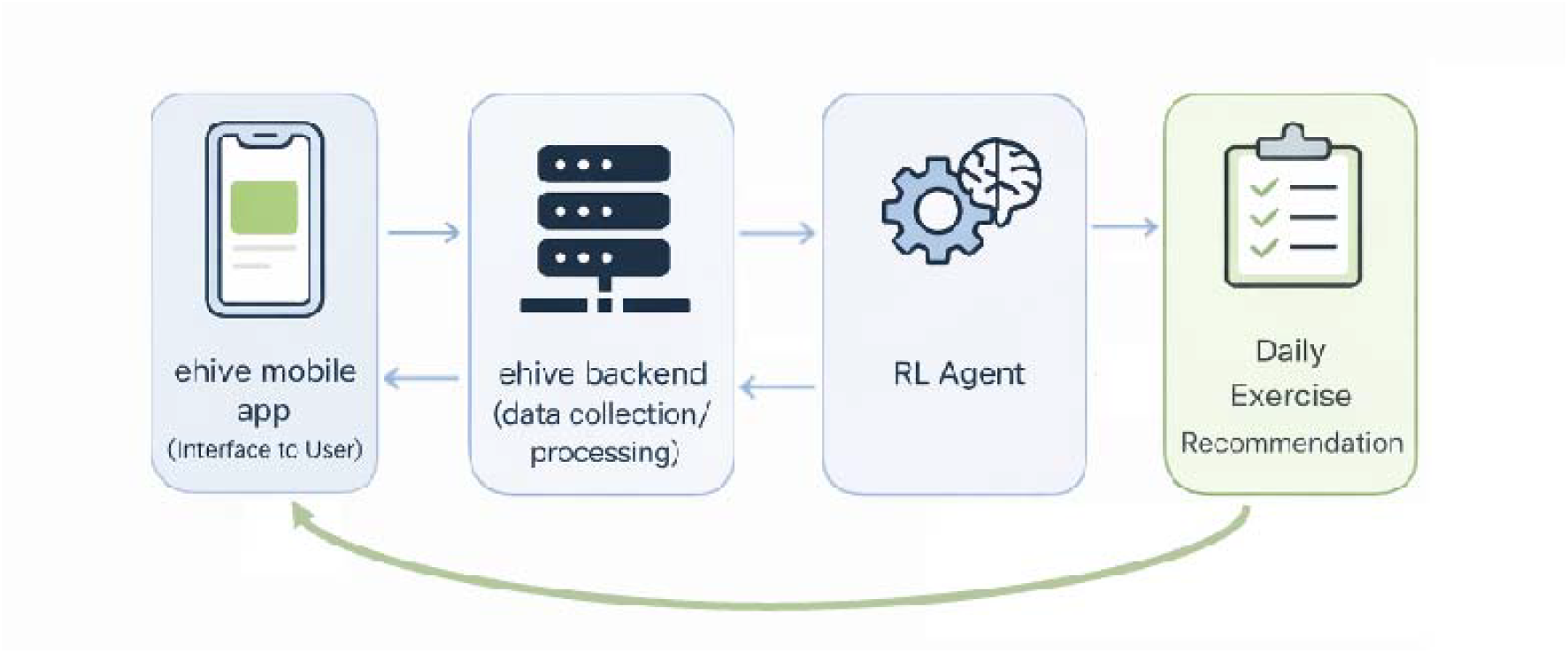
Schematic overview of the WorkoutCPP system architecture. The ehive mHealth platform integrates participant-reported symptom data and Fitbit wearable data to inform the contextual-bandit reinforcement learning agent, which generates personalized daily exercise recommendations delivered through the app interface.

**Figure 3.**
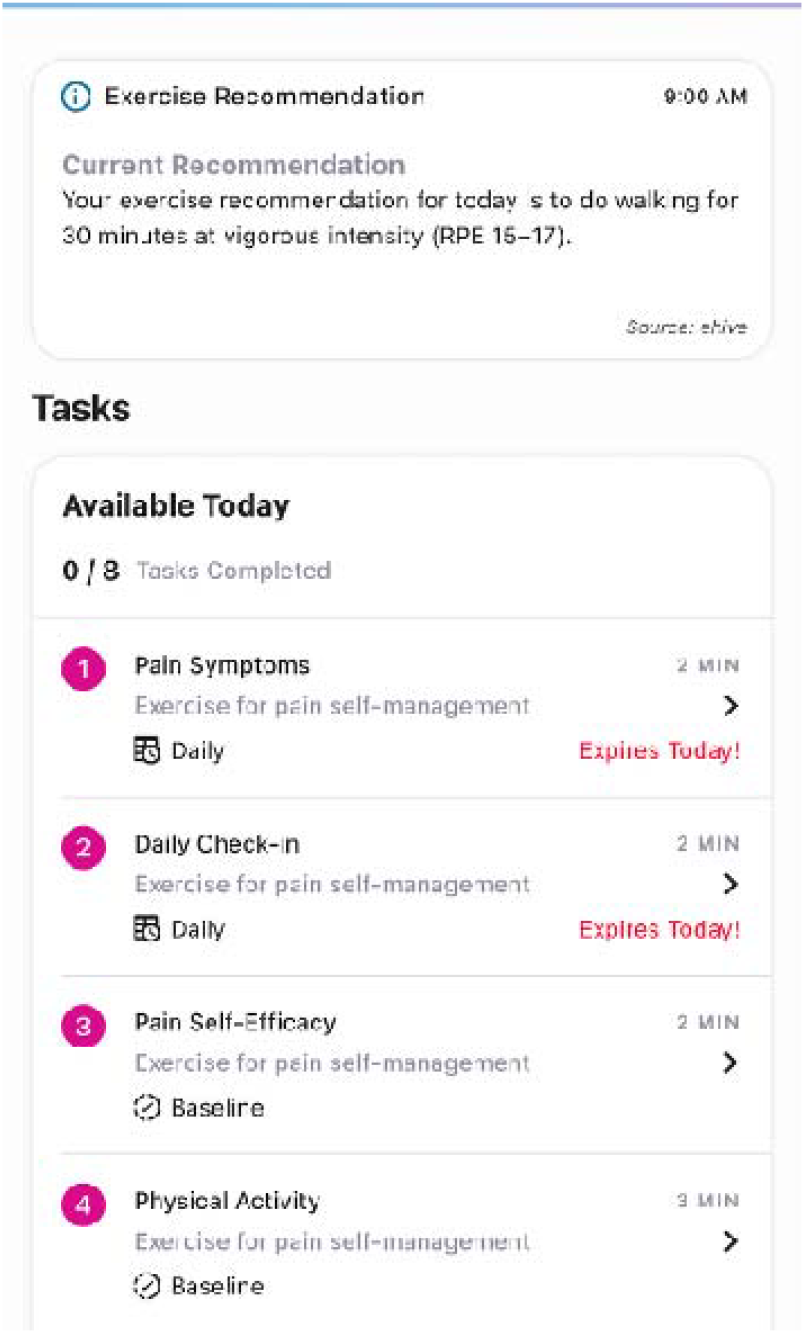
Screenshot of the *ehive* mobile application interface showing the exercise recommendation delivery and daily task completion features. The notification displays a sample RL-generated recommendation (top), while the task list shows daily and baseline self-tracking surveys completed by participants throughout the study, including pain symptoms, daily check-in, pain self-efficacy, and physical activity assessments.

All study procedures will be conducted through the ehive mobile-health platform developed at the Icahn School of Medicine at Mount Sinai, which integrates wearable device data and participant-reported information. Participants can attend an optional in-person onboarding visit and will complete the remaining study activities remotely.

### Participants and recruitment

Participants will be adults aged 18–55 years with a clinician or surgical diagnosis of a CPPD or symptoms consistent with chronic pelvic pain (e.g., dysmenorrhea, dyspareunia, dysuria, or bowel-associated pain persisting ≥1 year). Recruitment will occur through Mount Sinai clinical departments, including outreach via the OBGYN department and other affiliated clinics, the Mount Sinai Clinical Research Portal, and outreach to prior study participants.

### Inclusion criteria

1. Self-reported CPPD (e.g., endometriosis, adenomyosis, fibroids, etc.) based on clinician diagnosis
2. Aged 18–55 years.
3. Ownership of an iOS or Android smartphone.
4. Willingness to self-track daily symptoms, exercise activities, and self-management behaviors using the ehive app.
5. Willingness to wear an activity tracker for the study duration.
6. Willingness to follow exercise recommendations from a research smartphone App, provided no adverse symptoms occur.
7. Ability to read and write in English sufficient to understand study materials and communications.
8. At least intermittently physically active (e.g., ≥30 minutes of walking twice per week).

### Exclusion criteria

1. Absolute contraindications to PA (e.g., recent myocardial infarction, complete heart block, acute congestive heart failure, unstable angina, or uncontrolled severe hypertension, BP ≥180/110 mm Hg).
2. More than two “Yes” responses on the Physical Activity Readiness Questionnaire (PAR-Q) (17) without physician clearance.
3. Major life events expected during the next 10 weeks (e.g., pregnancy, planned surgery, or extended travel likely to interfere with participation).
4. Current or planned pregnancy within the next 6 months.
5. Having given birth in the past 6 months or currently nursing.
6. Inability to wear an activity tracker or use the ehive app for the study duration.
7. Complete inactivity (i.e., <60 minutes of moderate-intensity PA per week).

Screening and electronic informed consent will be completed through the ehive platform. Participants will receive $20 per week of active participation (up to $180 total) as compensation for time and data entry. Participants will be provided with Fitbit devices for passive exercise and PA data collection concurrent with their self-tracking in the ehive application.

To promote participant retention, daily push notifications are delivered through the ehive app to remind participants of their current exercise recommendation. Participants who fail to complete three daily check-ins within a given study week receive a compliance email from the study coordinator as an additional prompt. Weekly compensation of $20 serves as a further incentive for sustained engagement. In the event of participant withdrawal, all data collected prior to discontinuation will be retained for analysis provided the participant has completed at least one week in each study arm, as this represents the minimum threshold for informative within-person comparison. Data from participants who withdraw before meeting this threshold will be excluded from analysis, as insufficient exposure to both arms precludes meaningful estimation of individual treatment effects.

Participants are not required to modify any existing medications or treatments for CPPDs during the study period. Concomitant pharmacological and non-pharmacological pain management strategies are permitted throughout. The only functional requirement is that participants remain able to engage in regular physical activity from light to vigorous intensities; any new medical condition or treatment that precludes regular physical activity would constitute grounds for discontinuation as described above. Participants are asked to report any new medications or treatments initiated during the study through the ehive daily tracking survey to allow assessment of potential confounding.

### Intervention

The WorkoutCPP intervention delivers personalized exercise recommendations through the ehive app, informed by an embedded contextual-bandit RL agent. The RL agent uses a contextual bandit framework that updates the probability of selecting each recommendation based on observed participant responses, balancing exploration of new recommendations with exploitation of previously effective ones.

The features within the RL agent are described below:

- **Action set:** A set of exercise types encompassing all possible modalities (i.e., aerobic, strength, stretching, multi-modal, etc.) and intensities (i.e., light, moderate, vigorous). This action set is created for each participant, based on an intake survey they complete upon enrollment providing the types of activities they can perform and the equipment they have access to. This ensures that recommendations are restricted to feasible and safe options tailored to each participant’s capabilities and environment. An exercise physiologist reviews these responses and meets with the participant to verify their action set, provide exercise safety guidelines, and answer any questions the participant might have. This meeting takes approximately 30 minutes and is held virtually. The exercise physiologist on the study team (A.D.) holds an MS degree and is certified as an Exercise Physiologist (EP-C) by the American College of Sports Medicine (ACSM).
- **Context features:** Exercise type, intensity, and duration of the prior recommendation; the participant’s most recently reported composite pain score at the time of the decision; and running averages of the participant’s intensity and duration across prior exercise sessions. The composite pain score is calculated from severity ratings across all body locations the participant reports as painful that day, an approach that captures more granular, clinically meaningful pain information than a single global pain rating. This body-location detail informs the composite score but is not provided to the RL agent as a separate feature, given the limited sample size available for estimating location-specific effects.
- **Reward function:** Change in the participant’s composite pain score from the current day to the following day, *Pain*(*t*_*i*_ ) - *Pain*(*t*_*i*+1_), such that a decrease in next-day pain yields a positive reward. Adherence and adverse-symptom penalization are not currently incorporated into the reward calculation. These feedback points will be integrated manually into the RL agent’s behavior. If a participant reports a negative response, i.e., worsened pain or request to remove a recommendation, we will update the action set to remove that activity.
- **Safety and fallback behavior:** The system includes a fallback mechanism to ensure that a recommendation is always delivered when the RL model cannot produce a valid output, such as when there are insufficient data, non-informative participant activity histories, or computational timeouts. In these cases, the RL-generated probabilities are not used, and a random recommendation is instead selected from the participant’s predefined intervention set. These fallback recommendations are limited to activities and intensities that the participant has already indicated they are willing and able to perform during intake, ensuring that all delivered recommendations remain within a safe and pre-approved set. Furthermore, participants have the option to request the removal of an exercise if they experience any perceived unpleasant symptoms. Meanwhile, the RL model itself incorporates pain as a continuous predictor, which probabilistically reduces the likelihood of recommending higher-intensity or longer-duration activities when recent pain is elevated.

Participants may withdraw from the study at any time without penalty and will receive pro-rated compensation for completed weeks. The study team may discontinue a participant’s involvement if they report adverse events related to exercise that exceed expected minimal risk, develop new contraindications to physical activity, or become unreachable for an extended period. Data collected prior to withdrawal will be retained for analysis.

All adverse events will be reported to the Mount Sinai IRB within 48 hours per institutional policy.

Participants will receive one recommendation each day during every 2-week phase. The RL system will adapt recommendations in real time based on participant feedback and wearable data (Fitbit). Data collected during the one-week baseline period will be used to initialize the contextual model, calibrate individual context variables, and establish each participant’s symptom and activity history before the intervention period begins.

The exercise physiologist on the study team reviews the recommendations daily to monitor participant responses and ensure that recommendations remain safe and appropriate. Active comparator arm recommendations are standardized, evidence-based activities of comparable intensity and frequency, delivered without RL adaptation. Participants are randomized to the ABAB or BABA sequence in a 1:1 ratio using unconstrained simple randomization via an automated, computer-generated Python sequence without blocking or stratification, consistent with the within-person inference framework of the N-of-1 design. Given this framework, perfect balance across sequences is not required for valid estimation of individual treatment effects; sequence assignment primarily serves to control for order effects at the individual level. Any observed imbalance in sequence assignments reflects natural variation inherent to simple randomization under rolling enrollment and does not compromise the integrity of within-person comparisons.

To ensure strict allocation concealment, the random sequence is embedded within the secure backend software architecture and remains concealed from study personnel until the moment of assignment. The automated system handles both enrollment tracking and sequence assignment sequentially, ensuring that research personnel recruiting, screening, and enrolling participants have no prior access to or influence over the allocation sequence before the completion of the baseline week. Furthermore, participants are not informed of their sequence assignment at any point during the study, as knowledge of arm assignment could introduce expectation bias and compromise the integrity of within-person comparisons in the N-of-1 framework; participants receive daily recommendations through the ehive app without awareness of which study arm they are currently in. This study duration was selected to provide sufficient within-person observations across the full ABAB sequence while balancing participant burden and maintaining engagement, a design expected to yield up to approximately 2,835 person-level days of data for analysis.

Unblinding is not anticipated during the study. If a participant experiences an adverse event requiring clinical management, the study coordinator will disclose the participant’s current study arm assignment to the relevant clinical personnel as needed for their care. Participants who request to know their sequence assignment during the study period may be denied disclosure at the discretion of the PI, as unblinding could introduce expectation bias and compromise the integrity of within-person comparisons. Sequence assignments may be disclosed to participants upon study completion if requested.

### Study Variables

Study variables are listed in Table 1. They include participant demographics (assessed at baseline), symptoms, and contextual features used by the RL algorithm to generate adaptive exercise recommendations such as changes in reported pain between consecutive days and characteristics of prior exercises (e.g., intensity and duration). Data will be collected through the ehive application, with wrist-worn activity tracker data (e.g., Fitbit PA tracking) integrated into the platform for continuous monitoring.

**Table 1.**
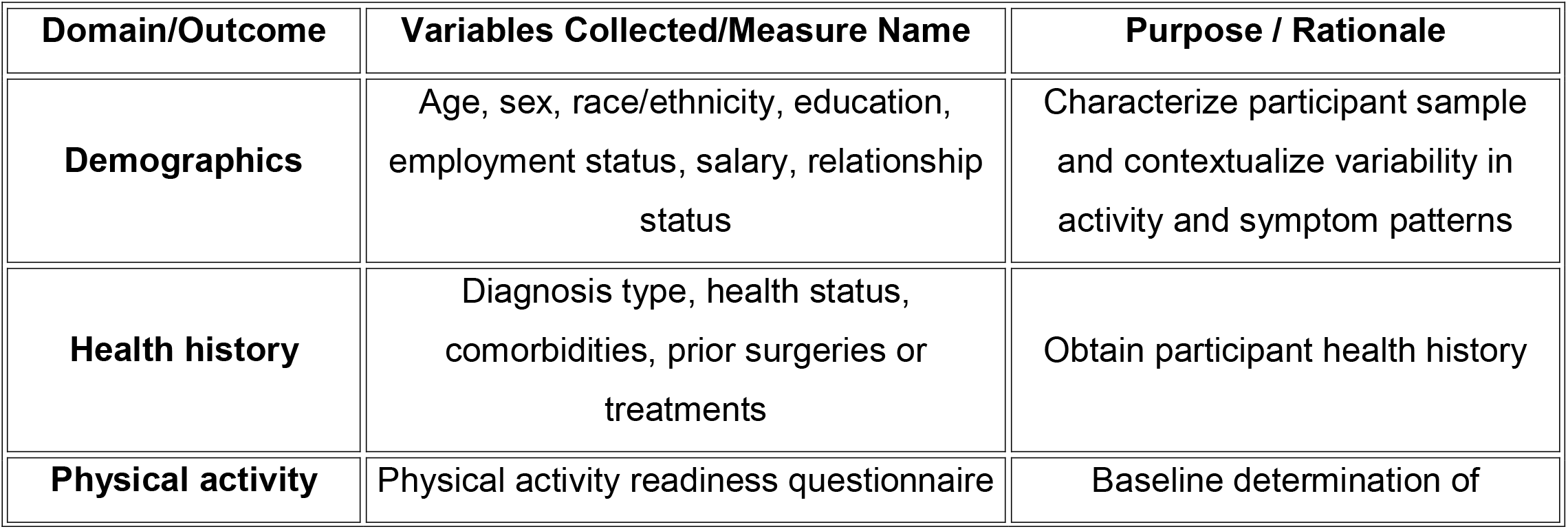
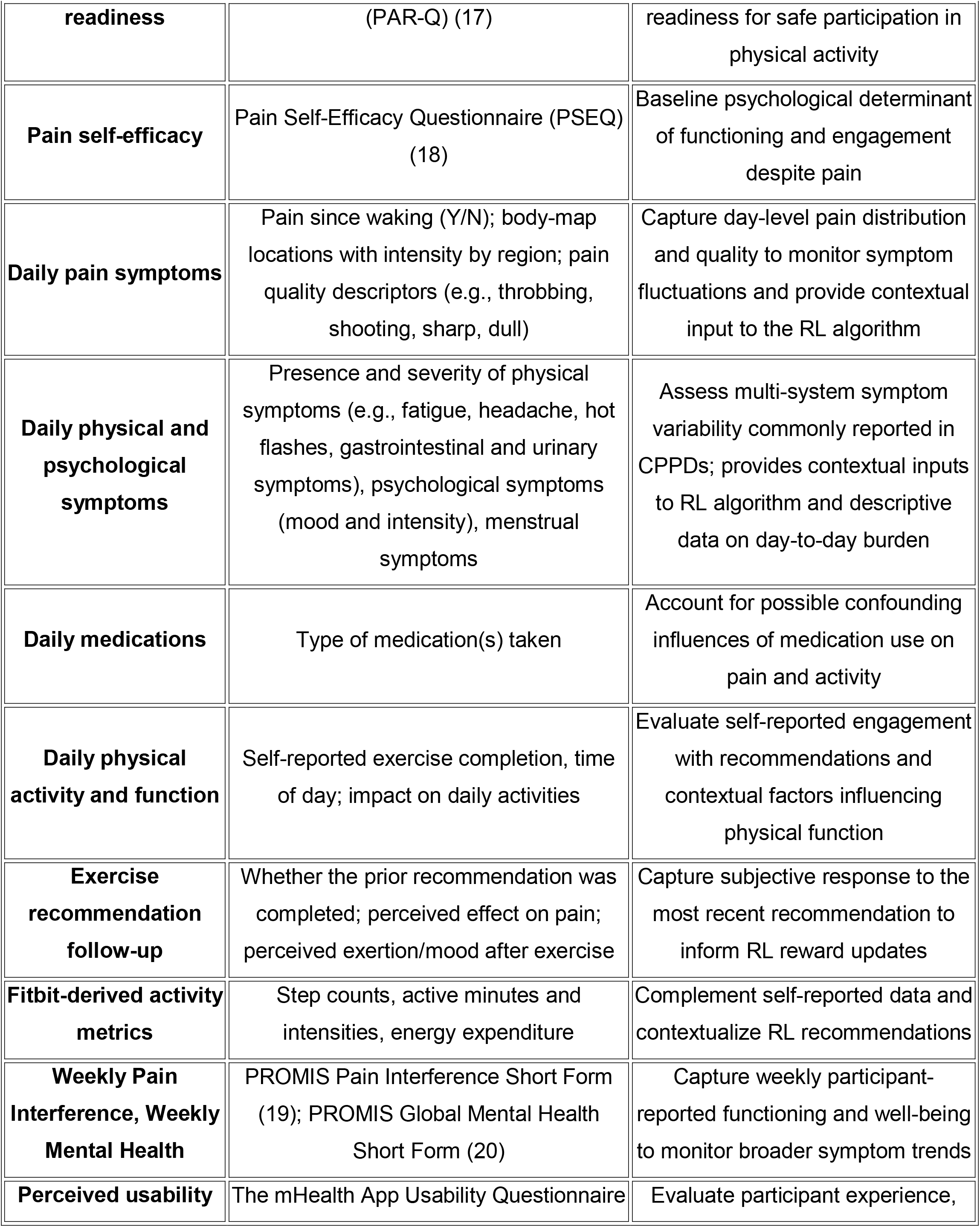
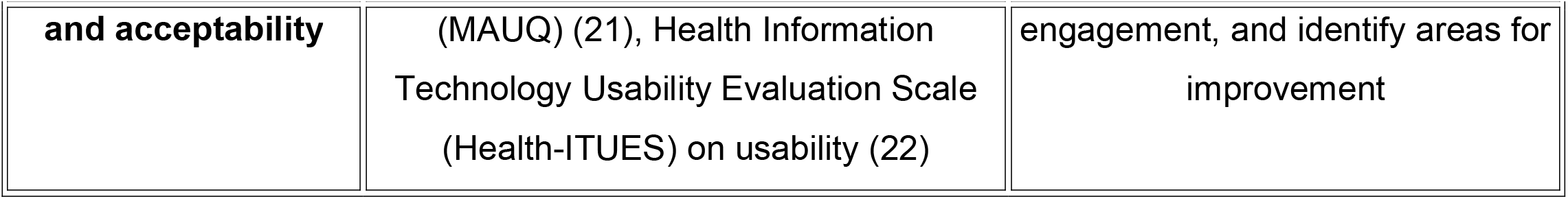
Overview of study variables collected in the *WorkoutCPP* pilot N-of-1 trials across baseline, daily, weekly, and post-intervention timepoints, including Fitbit wearable data. Surveys are administered through the *ehive* mobile application.

| Domain/Outcome | Variables Collected/Measure Name | Purpose / Rationale |
| --- | --- | --- |
| Demographics | Age, sex, race/ethnicity, education, employment status, salary, relationship status | Characterize participant sample and contextualize variability in activity and symptom patterns |
| Health history | Diagnosis type, health status, comorbidities, prior surgeries or treatments | Obtain participant health history |
| Physical activity | Physical activity readiness questionnaire | Baseline determination of |

| **Domain/Outcome** | **Variables Collected/Measure Name** | **Purpose / Rationale** |
| --- | --- | --- |
| **readiness** | (PAR-Q) (17) | readiness for safe participation in physical activity |
| **Pain self-efficacy** | Pain Self-Efficacy Questionnaire (PSEQ) (18) | Baseline psychological determinant of functioning and engagement despite pain |
| **Daily pain symptoms** | Pain since waking (Y/N); body-map locations with intensity by region; pain quality descriptors (e.g., throbbing, shooting, sharp, dull) | Capture day-level pain distribution and quality to monitor symptom fluctuations and provide contextual input to the RL algorithm |
| **Daily physical and psychological symptoms** | Presence and severity of physical symptoms (e.g., fatigue, headache, hot flashes, gastrointestinal and urinary symptoms), psychological symptoms (mood and intensity), menstrual symptoms | Assess multi-system symptom variability commonly reported in CPPDs; provides contextual inputs to RL algorithm and descriptive data on day-to-day burden |
| **Daily medications** | Type of medication(s) taken | Account for possible confounding influences of medication use on pain and activity |
| **Daily physical activity and function** | Self-reported exercise completion, time of day; impact on daily activities | Evaluate self-reported engagement with recommendations and contextual factors influencing physical function |
| **Exercise recommendation follow-up** | Whether the prior recommendation was completed; perceived effect on pain; perceived exertion/mood after exercise | Capture subjective response to the most recent recommendation to inform RL reward updates |
| **Fitbit-derived activity metrics** | Step counts, active minutes and intensities, energy expenditure | Complement self-reported data and contextualize RL recommendations |
| **Weekly Pain Interference, Weekly Mental Health** | PROMIS Pain Interference Short Form (19); PROMIS Global Mental Health Short Form (20) | Capture weekly participant-reported functioning and well-being to monitor broader symptom trends |
| **Perceived usability** | The mHealth App Usability Questionnaire | Evaluate participant experience, |
| and acceptability | (MAUQ) (21), Health Information Technology Usability Evaluation Scale (Health-ITUES) on usability (22) | engagement, and identify areas for improvement |

### Primary outcomes

This study has two primary outcomes focused on feasibility: exercise recommendation adherence rate and participant retention rate, both assessed at 9 weeks at study completion per participant. Exercise recommendation adherence rate is calculated as the proportion of daily exercise recommendations completed over the course of the intervention, with a higher rate indicating that participants are completing their given exercise recommendations at higher frequencies. Participant retention rate is the proportion of enrolled participants completing study participation until the end of intervention, with a higher rate indicating that participants complete the 9-week intervention period at higher frequencies.

These two outcomes will be evaluated alongside additional feasibility indicators, including participant enrollment and completeness of daily symptom and activity tracking, to assess whether the ehive platform and RL-driven workflow are practical for sustained use in individuals with CPPDs. These broader feasibility indices will also inform planning for subsequent trials, including adjustments to account for participant attrition and variability in adherence.(23, 24)

Within evaluating feasibility, we will monitor the safety of the study through the number and proportion of participants reporting adverse events (AEs) related to exercise, with events monitored and reviewed weekly by the study team. Light-to moderate-intensity activity interventions are typically considered low risk in chronic pain populations, but systematic AE monitoring will ensure participant protection and compliance with institutional ethics requirements. (9, 10)

### Secondary outcomes

The interpretability of the RL algorithm will be assessed using entropy of the agent’s action probability distributions, calculated at each decision point throughout the study period. Entropy ranges from a minimum of 0, indicating the agent selects a single action with certainty, to a maximum of log(K), where K is the number of distinct recommendation actions available at that decision point, representing equal probability assigned across all available actions. Decreasing entropy over time will indicate increasing model confidence and more consistent personalized recommendation patterns, while higher entropy indicates greater uncertainty in the agent’s decision making. This outcome will be evaluated across the full duration of the intervention using daily data. Such measures align with published recommendations for evaluating explainable and safe AI in digital health applications. (23, 24)

### Exploratory outcomes

#### Contextual sensitivity

Contextual sensitivity will be examined as variability in the RL agent’s action probability distributions across differing daily symptom states, assessed separately for pain severity, fatigue, and mood as three distinct contextual dimensions. Variability will be quantified using total variation distance between action probability distributions across distinct symptom-state contexts, ranging from a minimum of 0, indicating identical action probability distributions across symptom states and no contextual sensitivity, to a maximum of 1, indicating completely non-overlapping distributions. Greater total variation distance indicates higher contextual sensitivity, reflecting more responsive adaptation of exercise recommendations to individual symptom fluctuations. This measure is assessed from logged probability distributions generated at each daily decision point and evaluated across the full duration of the intervention.

As a complementary characterization of the agent’s behavior over time, action probabilities will also be examined using time-series analysis to assess how recommendation preferences evolve across the intervention, including convergence toward stable recommendations or continued exploration, and fallback event rates will be monitored to assess how often safety constraints override RL-generated recommendations. (12, 13) However, observed patterns may also be influenced by the limited amount of data available during early study phases and by model priors, which may affect the rate and extent of adaptation.

#### Symptom and behavioral changes

Daily symptom reports, activity data, and weekly patient-reported outcomes will be examined to characterize individual trajectories and patterns of engagement between the two study arms. This includes change in daily pain severity, measured as the difference between a participant’s mean pain intensity rating on a given day and the following day (*Pain*(*t*_*i*_ ) - *Pain*(*t*_*i*+1_)), calculated from severity ratings across all body locations the participant identifies as painful that day. A positive value indicates reduced next-day pain severity; a negative value indicates worsening. This measure is also used by the RL agent in calculating reward. Analyses will primarily focus on within-person comparisons, with secondary summaries describing trends across participants.

#### Perceived usability and acceptability

At study completion, participants will complete the MAUQ (21) and the Health-ITUES (22) to assess perceived usability, satisfaction, and perceived helpfulness of the adaptive recommendations, to evaluate participant experience and guide refinement of digital behavioral interventions in line with published recommendations.(21, 22)

### Sample size

As a pilot feasibility study, formal power calculations were not conducted. The target enrollment of 45 participants anticipates an attrition rate of ≈ 33%, yielding approximately 30 completers. This sample size is expected to provide sufficient precision to estimate feasibility metrics and to characterize within-person variability needed for algorithm refinement and future power calculations.

### Data Management and Monitoring

All study data, including participant histories, recommendations, and RL probability logs, will be stored on secure Mount Sinai servers with encrypted transmission and automatic daily backups. Participant data will be pseudonymized at collection using unique ehive-generated identifiers, and linkage keys connecting these identifiers to personally identifiable information will be stored separately and securely.

Study monitoring will include both automated and manual quality assurance procedures. Automated alerts will flag anomalies in real time (e.g., missing recommendations, repeated high-pain reports, crashes in backend study processes). In addition, the study team will conduct routine monitoring that includes a twice-daily review of incoming symptom reports, daily summaries of participant activity and symptom trends, and weekly summaries of adherence and data completeness. Monitoring procedures will also include verification that recommendations are delivered as scheduled and onboarding checks to ensure that newly enrolled participants are correctly integrated into the system workflow. Only authorized study personnel approved by the Mount Sinai Institutional Review Board will have access to identifiable information, while all analyses will be conducted using pseudonymized data. Overall trial conduct is reviewed on a weekly basis by the lead investigator (BTT) and discussed with the PI (IE) at regular check-in meetings to ensure ongoing protocol adherence and timely resolution of any operational issues.

No independent data monitoring committee has been established given the minimal risk profile and pilot scope of the study; safety oversight is provided by the study team as described above. No formal interim efficacy analyses with pre-specified stopping rules are planned. Ongoing operational monitoring of RL system performance and preliminary data trends will be conducted throughout the study to ensure system integrity and participant safety, as described above. The study may be stopped early in the event of a pattern of serious adverse events related to exercise or a system failure that compromises participant safety or data integrity.

### Data analysis plan

#### Primary Feasibility and Safety Outcomes

Exercise recommendation adherence rate and participant retention rate will be summarized as proportions with 95% confidence intervals (CIs). We will evaluate system safety by computing the proportion of participants reporting exercise-related adverse events. Feasibility estimates will be compared against pre-specified benchmark thresholds (≥70% adherence rate; ≥60% data completeness) to inform future trial power calculations.

Preliminary treatment effects and algorithmic analysis. For all comparative symptom analyses, algorithm interpretability metrics, and contextual sensitivity modeling, we will include participants who complete the 1-week baseline period and at least 1 full week in each study arm. Because N-of-1 trials evaluate individual-level intervention effects across alternating blocks, participants who discontinue prior to completing at least one period per arm do not yield sufficient cross-arm exposure to compute within-person treatment estimates, motivating the decision to follow a modified intent-to-treat (mITT) approach.

#### Algorithm performance and interpretability

Entropy of the RL agent’s action probability distributions will be summarized descriptively over time, with decreasing entropy interpreted as increasing model confidence. Trends will be examined across the full intervention period using daily data.

#### Contextual sensitivity

Contextual sensitivity will be analyzed by comparing the RL agent’s action probability distributions across differing daily pain, fatigue, and mood states, using total variation distance between these distributions as the measure of how responsively the model adapts to participants’ symptom states. To characterize the agent’s behavior, action probabilities will also be examined using time-series analyses to assess how recommendation preferences evolve over the course of the intervention, including convergence toward stable recommendations or continued exploration. We will monitor fallback event rates to assess how often safety constraints override recommendations.

#### Symptom and behavioral changes

Daily and weekly patient-reported outcomes, including pain, fatigue, mood, and activity, will be analyzed using statistical models designed for repeated within-person measurements over time. These models will estimate how each participant’s outcomes changed between the active comparator and experimental study arms, allowing both participant-specific comparisons and a summary of overall patterns across the sample. Analyses will primarily emphasize within-person effects, consistent with the N-of-1 design, with population-level estimates treated as descriptive summaries.

#### Perceived usability and acceptability

Scores on the MAUQ and Health-ITUES will be summarized descriptively to characterize perceived usability, satisfaction, and helpfulness of the adaptive recommendations.

#### Missing data

Missing daily self-report data will be evaluated for patterns of missingness and addressed using approaches appropriate for longitudinal data with repeated measures. Where appropriate, we will conduct sensitivity analyses to evaluate the potential impact of missing data on study findings, including the stability of feasibility and treatment estimates across varied minimum adherence thresholds.

Sensitivity analyses will examine the robustness of feasibility estimates under different adherence thresholds and missing data assumptions. Exploratory subgroup analyses may be conducted by diagnosis type (e.g., endometriosis vs. other CPPDs) and randomization sequence (ABAB vs. BABA) to assess potential order effects and will be interpreted descriptively given the pilot nature and limited sample size of the study.

## Supporting information

SPIRIT 2025 Checklist

## Ethics and Dissemination

Ethical approval for this study was obtained from the Institutional Review Board at the Icahn School of Medicine at Mount Sinai (Protocol # STUDY-23-00721). All study procedures will adhere to the principles of the Declaration of Helsinki and International Council on Harmonisation Good Clinical Practice (ICH-GCP) guidelines. Written informed consent will be obtained electronically through the ehive platform before any study activity. Participants may withdraw from the study at any time without penalty or loss of compensation for completed activities. No study procedures have occurred prior to IRB approval. Any important modifications to the protocol will be submitted to the Mount Sinai IRB for approval prior to implementation and communicated to all active participants if the modification affects their participation.

The anticipated risks of this behavioral mobile health intervention are minimal and primarily limited to light physical exertion. To mitigate these risks, exercise recommendations are tailored dynamically to individual physical abilities and symptom status; furthermore, the reinforcement learning (RL) algorithm features predefined safety constraints, including probability caps and fallback options, to prevent the delivery of overly strenuous activities. Adverse events will be documented, reviewed weekly by the research team, and reported to the IRB in accordance with institutional policy. Because of this low-risk, non-invasive design, a formal independent Data Monitoring Committee (DMC) was deemed unnecessary, with continuous safety and data oversight managed directly by the principal investigators and the institutional review board. If a participant believes that participation in this study has caused them harm, they should contact the Lead Researcher. Participants who withdraw from the study, including due to an adverse event, will receive pro-rated compensation for weeks of participation already completed, consistent with the compensation terms described above.

Confidentiality will be protected by storing all data on secure, encrypted Mount Sinai servers with restricted access. Data will be de-identified at collection, and linkage keys will be stored separately. No personally identifiable information will appear in analyses or publications.

Study findings will be disseminated through peer-reviewed publications (including this protocol and subsequent outcome papers), conference presentations, and de-identified summary reports shared with participants and collaborators. Individual participant data will not be shared, in order to protect participant privacy and confidentiality, consistent with the informed consent form under which participants were enrolled. Results from this pilot will inform refinement of the adaptive intervention and the design of a future randomized controlled trial. This study protocol was registered on the Open Science Framework (OSF Registries; https://osf.io/d9f36) prior to participant enrollment. Trial registration on ClinicalTrials.gov was not initiated at the time of study launch, as the pilot was not originally designed to evaluate clinical efficacy. Recognizing the value of prospective transparency for adaptive intervention trials, the study was subsequently registered on ClinicalTrials.gov (NCT07810218) retrospectively, following the recommendations of the ICMJE for trials informing future confirmatory research.

## Data Availability Statement

This is a protocol paper describing an ongoing pilot study. Data collection is currently in progress. Analytical code will be made available upon reasonable request following study completion, subject to institutional data governance policies. Individual participant data will not be shared, consistent with the informed consent form under which participants were enrolled. The study protocol and companion SPIRIT checklist are registered on the Open Science Framework (https://osf.io/d9f36). All relevant protocol materials, including final versions of the manuscript and compliance checklists, are publicly accessible via the OSF project repository files.

## Protocol Availability

This manuscript constitutes the study protocol. The statistical analytic code can made available upon reasonable request upon completion of all data analyses. No supplementary protocol document exists beyond this publication.

## Patient and Public Involvement

Patients or members of the public were not formally involved in the design, conduct, reporting, recruitment, or dissemination of plans for this research. Participants were not asked to assess the burden of the intervention and time required to participate in the study. Study findings will be disseminated with participants and the broader community through plain language summaries.

## Author Contributions

IE and SK conceptualized the overall study design, provided supervision throughout the project, and obtained the funding. MD and KL developed the ehive application infrastructure, backend architecture, and supported system integration and troubleshooting. SK and JV provided guidance on the RL framework. BTT is responsible for the development of the RL framework and its integration with ehive, ongoing monitoring of the study backend and system performance, with support from JV and RC. CO, AK, JR, and GO managed participant-facing study operations, including weekly adherence tracking, participant communication, compensation distribution, and maintenance of study data in REDCap (Research Electronic Data Capture). AD serves as the exercise physiologist providing exercise safety and adherence guidance to the participants. SH and SKh provided clinical expertise to inform study design. BTT drafted the initial manuscript with guidance from IE and SK. All authors contributed to the manuscript for important intellectual content, approved the final version for submission, and agree to be accountable for all aspects of the work.

